# Impact of Population Mixing Between a Vaccinated Majority and Unvaccinated Minority on Disease Dynamics: Implications for SARS-CoV-2

**DOI:** 10.1101/2021.12.14.21267742

**Authors:** David N. Fisman, Afia Amoako, Ashleigh R. Tuite

## Abstract

**Background:** The speed of vaccine development has been a singular achievement during the SARS-CoV-2 pandemic, though uptake has not been universal. Vaccine opponents often frame their opposition in terms of the rights of the unvaccinated. Our objective was to explore the impact of mixing of vaccinated and unvaccinated populations on risk among vaccinated individuals.

**Methods:** We constructed a simple Susceptible-Infectious-Recovered (SIR) compartmental model of a respiratory infectious disease with two connected sub-populations: vaccinated individuals and unvaccinated individuals. We simulated a spectrum of patterns of mixing between vaccinated and unvaccinated groups that ranged from random mixing to like-with-like mixing (complete assortativity), where individuals preferentially have contact with others with the same vaccination status. We evaluated the dynamics of an epidemic within each subgroup, and in the population as a whole.

**Results:** The relative risk of infection was markedly higher among unvaccinated individuals than among vaccinated individuals. However, the contact-adjusted contribution of unvaccinated individuals to infection risk during the epidemic was disproportionate, with unvaccinated individuals contributing to infections among the vaccinated at a rate higher than would have been expected based on contact numbers alone. As assortativity increased, attack rates among the vaccinated decreased, but the contact-adjusted contribution to risk among vaccinated individuals derived from contact with unvaccinated individuals increased.

**Interpretation:** While risk associated with avoiding vaccination during a virulent pandemic accrues chiefly to the unvaccinated, the choices of unvaccinated individuals impact the health and safety of vaccinated individuals in a manner disproportionate to the fraction of unvaccinated individuals in the population.

## Introduction

The remarkable speed of vaccine development, production and administration is a singular human achievement during the SARS-CoV-2 pandemic (1). While the ability to vaccinate to herd immunity has been held back by the increasing transmissibility of novel variants of concern (e.g., Delta and Omicron variants) (2, 3), and global distribution of vaccines is deeply inequitable (4), the effectiveness of vaccines against acquisition of infection, in reducing severity of disease, and in disrupting onward transmission even when breakthrough infections occur, is likely to have saved many lives. The recent emergence of the immune-evasive Omicron variant may undermine some of these gains, though provision of booster vaccine doses may restore vaccination to a high level of potency, and Omicron-tuned vaccines may emerge in 2022 (3, 5-7).

However anti-vaccine sentiment, fueled in part by organized disinformation efforts, has resulted in suboptimal uptake of readily available vaccines in many countries, with adverse health and economic consequences (8-10). Although the decision to adopt vaccination is often framed in terms of the rights of individuals to opt out of such programs (11, 12), such arguments neglect the potential harms to the wider community that derive from poor vaccine uptake. Non-vaccination is expected to result in amplification of disease transmission in unvaccinated sub-populations, but the communicable nature of infectious diseases means that this creates risk for vaccinated populations as well, when vaccines are imperfect. While assortative (like-with-like) mixing (13) is characteristic of many communicable disease systems, and this may be expected to limit interaction between vaccinated and unvaccinated sub-populations to some degree, the normal functioning of society means that complete assortativity is not a phenomenon observed in reality. Furthermore, the dominant airborne mode of spread of SARS-CoV-2 (14-20) means that close-range physical mixing of individuals from vaccinated and unvaccinated groups is not necessary for between-group disease transmission. Historically, behaviors that create health risks for the community as well as individuals have been the subject of public health regulation. This is true of communicable infectious diseases, but also applies to public health statutes that limit indoor cigarette smoking (21), and legal restrictions on driving under the influence of alcohol and other intoxicants (22, 23).

Simple mathematical models can often provide important insights into the behavior of complex communicable disease systems (13, 24, 25). To better understand the implications of the interplay between vaccinated and unvaccinated populations under different assumptions about population mixing, we constructed a simple Susceptible-Infectious-Recovered (SIR) model to reproduce the dynamics of interactions between vaccinated and unvaccinated population subgroups in a highly vaccinated population. Our objectives were: (i) to contrast contribution to epidemic size and risk estimates by sub-population; and (ii) to understand the impact of mixing between vaccinated and unvaccinated groups on expected disease dynamics.

## Methods

### Model

We constructed a simple compartmental model of a respiratory viral disease, as in (26). The model is described in detail in the attached **Technical Appendix**. The model represents individuals as residing in three possible “compartments”: susceptible to infection (S), infected and infectious (I), and recovered from infection with immunity (R). The compartments are divided to reflect two connected sub-populations: vaccinated individuals and unvaccinated individuals. Susceptible individuals move into the infectious compartment following effective contacts (i.e., contacts of a nature and duration sufficient to permit transmission) with infected individuals. In the context of an airborne virus like SARS-CoV-2 (14-20), effective contact may be conceptualized as “sharing air” with an infective case. Following an infectious period, infectious individuals recover with immunity. We also assumed that some fraction of the unvaccinated population had immunity at baseline due to prior infection and that a fraction of the population was vaccinated. Immunity following vaccination was treated as an all or none phenomenon, with only a fraction of vaccinated individuals (as defined by vaccine effectiveness) entering the model in the immune state and the remainder left in the susceptible state. For example, an 80% efficacious vaccine would result in 80% of vaccinated individuals becoming immune, with the remaining 20% susceptible to infection. We did not model waning immunity.

Human populations do not mix randomly and exhibit a tendency for individuals to interact preferentially with others like themselves (13, 27), a phenomenon is referred to as “assortativity”. The relative frequency of interactions between individuals within different groups occurs on a spectrum that lies between complete assortativity (i.e., exclusive like-with-like mixing) and random mixing. For instance, age assortative mixing is frequently observed: children are more likely to interact with other children than would be expected if contacts occurred at random across all age groups. The use of matrices to govern such interactions are described in detail in the **Technical Appendix**.

However, with respect to contacts between individuals from two different groups, relative frequency of contacts will depend both on the relative sizes of the two groups, and the degree of assortativity present. In our model, assortativity is determined by a constant, denoted η, with random mixing occurring when η = 0, complete assortativity occurring when η = 1, and intermediate degrees of assortativity occurring at other intermediate values. For our model, with 20% of the population unvaccinated, when random mixing is assumed (η = 0), 20% of the contacts a vaccinated person has would be expected to occur with unvaccinated people. When exclusively like-with-like mixing is assumed (η = 1), 0% of contacts a vaccinated person has would be with unvaccinated people. For intermediate levels of assortativity (η = 0.5), 10% of a vaccinated person’s contacts would be with unvaccinated people.

Our base case model was otherwise parameterized to represent a disease similar to SARS-CoV-2 infection with Delta variant, with an R_0_ (the reproduction number of an infectious disease in the absence of immunity or control) of 6 (28) and higher values were used to capture the dynamics of the Omicron variant (29). Our lower bound estimate for vaccine effectiveness (40%) reflected uncertainty regarding the emerging Omicron variant (3, 7), while our upper bound (80%) reflected the higher effectiveness seen with the Delta variant (30). We used the model to explore the impact of varying rates of immunization and different levels of assortativity on the dynamics of disease in vaccinated and unvaccinated sub-populations. We evaluated the absolute contribution to overall case counts by these sub-populations, as well as within-group and overall infection risk. Attack rates were calculated as the cumulative number of infections divided by the population size. We calculate a quantity that we denote ψ which we define as the cumulative incidence of infections among the vaccinated that derive from contact with unvaccinated individuals, divided by the fraction of the population that is unvaccinated. A version of the model in Microsoft Excel is available at <u>10.6084/m9.figshare.15189576</u>.

## Results

Simulated epidemics assuming different amounts of mixing between vaccinated and unvaccinated groups are presented in **Figure 1**. With 20% baseline immunity among unvaccinated individuals, and 80% of the population vaccinated, the absolute number of cases from vaccinated and unvaccinated groups was similar when mixing was random; however, after adjustment for substantially larger population in the vaccinated group, risk of infection was markedly higher among unvaccinated individuals during the epidemic. With increased like-with-like mixing, differences in incidence between the vaccinated and unvaccinated groups became more apparent, with cases in the unvaccinated population accounting for a substantial proportion of infections during the epidemic wave. Assortativity uncoupled the dynamics of vaccinated and unvaccinated populations, with unvaccinated populations experiencing higher and earlier peak incidence than vaccinated populations. Cumulative attack rates among the vaccinated were highest with random mixing and lowest with highly assortative mixing. By contrast, cumulative attack rates were lowest among the unvaccinated with random mixing, and highest with highly assortative mixing. The highest cumulative attack rates in the population overall were seen with intermediate levels of assortativity, although the differences were small.

**Figure 1.**
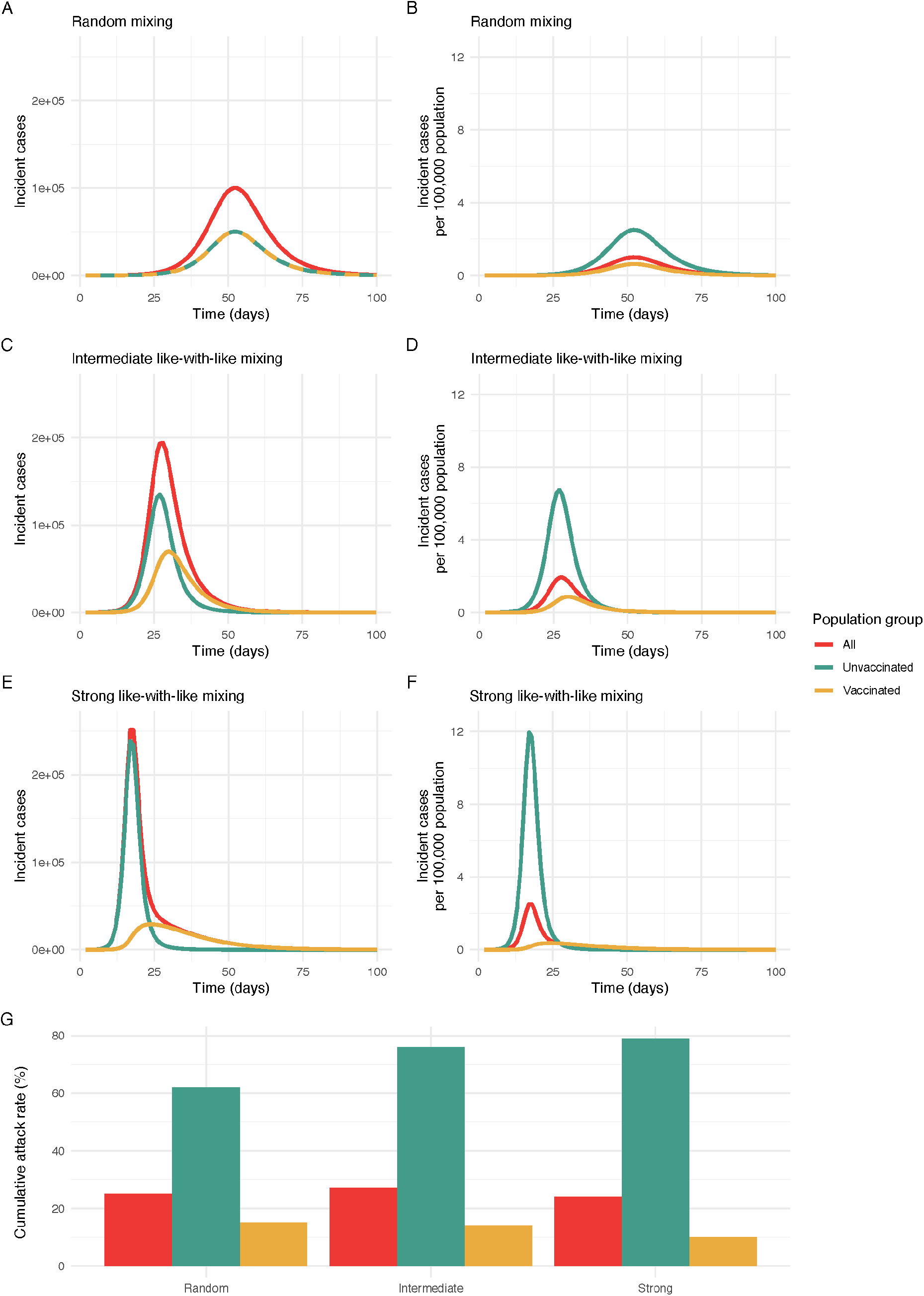
Simulated epidemics for different levels of mixing between vaccinated and unvaccinated populations. Incident cases (A, C, E) and population-adjusted incidence per 100,000 (B, D, F) in the unvaccinated, vaccinated, and overall modelled populations. The degree of assortativity (like-with-like mixing, η) varies from (A, B) random mixing (η=0) (random mixing), to (C, D) intermediate like-with-like mixing (η = 0.5), to (E, F) near exclusive mixing with people of the same vaccination status (η = 0.9). As like-with-like mixing increases, epidemic size among the vaccinated is smaller in absolute terms than among the unvaccinated, and also has a very different contour. Increasing assortativity increased cumulative attack rates among unvaccinated individuals, and decreased cumulative attack rates among vaccinated individuals (G); highest overall attack rates were seen with intermediate levels of assortativity.

Varying the degree of like-with-like mixing resulted in changes in epidemic size in the vaccinated population. As assortativity increased (i.e., with reduced contact between vaccinated and unvaccinated populations) the final attack rate decreased among vaccinated individuals, but the contribution of risk to vaccinated individuals due to infection acquired from contact with unvaccinated individuals (as measured by ψ, the ratio of the fraction of total infections derived from unvaccinated individuals to the proportion of contacts with unvaccinated individuals) increased. The larger the value of ψ, the more unvaccinated individuals contribute to infections in the vaccinated population.

This pattern was consistent across a range of values for vaccine effectiveness and reproduction numbers (**Figure 2**). Increased like-with-like mixing reduced final outbreak size among the vaccinated most markedly at lower reproduction numbers, but increased the value of ψ. With lower vaccine effectiveness, as is observed with the Omicron variant, the effects of assortativity were attenuated. With either lower reproduction numbers, or higher vaccine efficacy, transmission was more readily disrupted within the vaccinated subpopulation, such that risk arose increasingly from interactions with the unvaccinated subpopulation, where transmission continued. As assortativity increased, contribution to infection risk among the vaccinated was increasingly derived from (less and less common) interactions with unvaccinated individuals, increasing the value of ψ. Similar patterns were seen in sensitivity analyses in which vaccine coverage was increased from 80 to 99% (**Figure 3**). Increasing population vaccination coverage decreased the attack rate among vaccinated individuals (as expected, due to indirect protective effects) but further increased the relative contribution to risk in vaccinated individuals by the unvaccinated at any level of assortativity.

**Figure 2.**
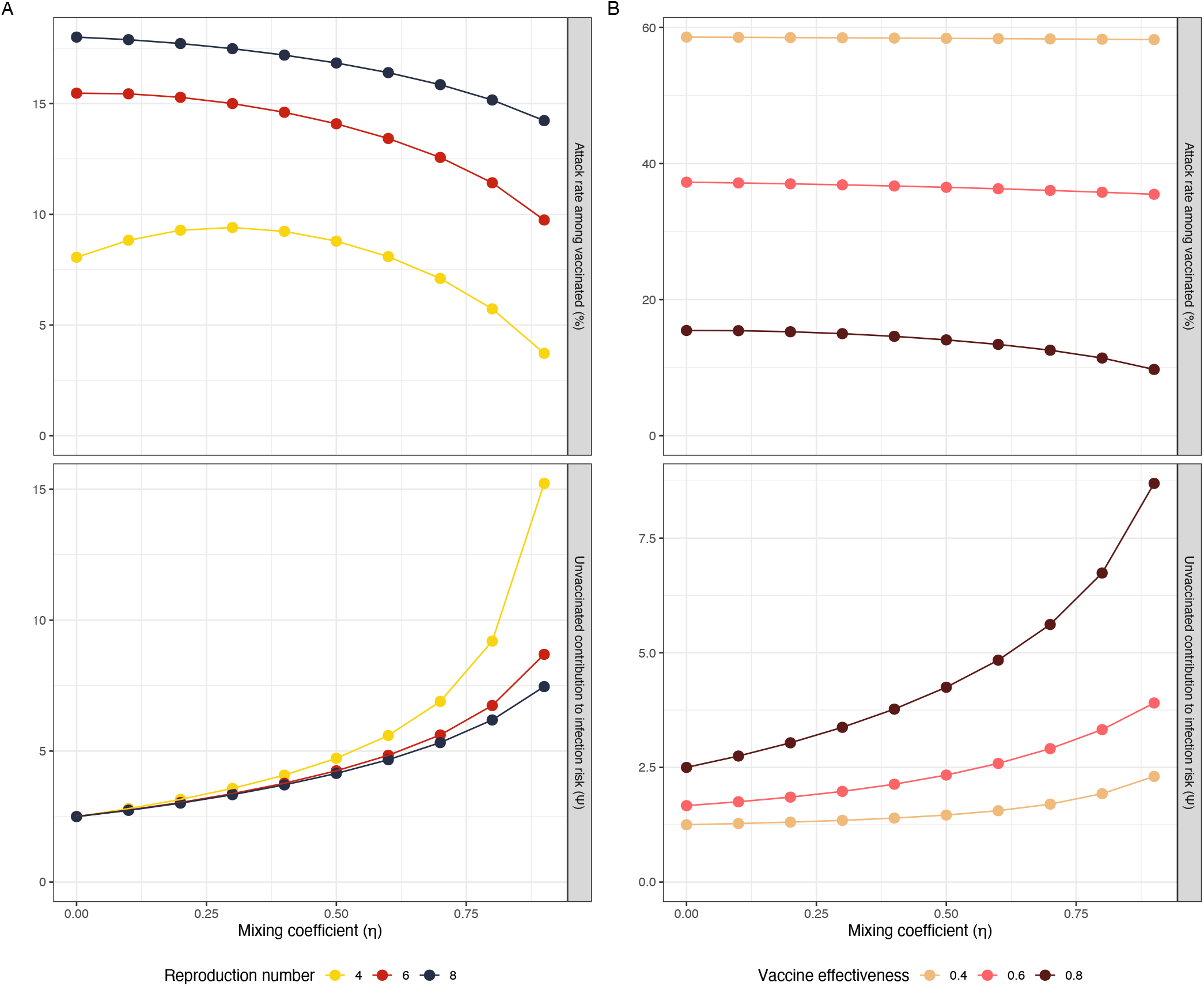
Impact of mixing between vaccinated and unvaccinated populations on contribution to risk and final epidemic size for varying reproduction numbers and vaccine effectiveness. Both panels demonstrate the impact of increasing like-with-like mixing on outbreak size among the vaccinated and contact-adjusted contribution to infection risk in the vaccinated by unvaccinated individuals (ψ). As like-with-like mixing (η) increases, the attack rate among vaccinated individuals decreases, but ψ increases. This relationship is seen across a range of (A) initial reproduction numbers and (B) vaccine effectiveness. These effects are more marked at lower reproduction numbers, and are attenuated as vaccines become less effective.

**Figure 3.**
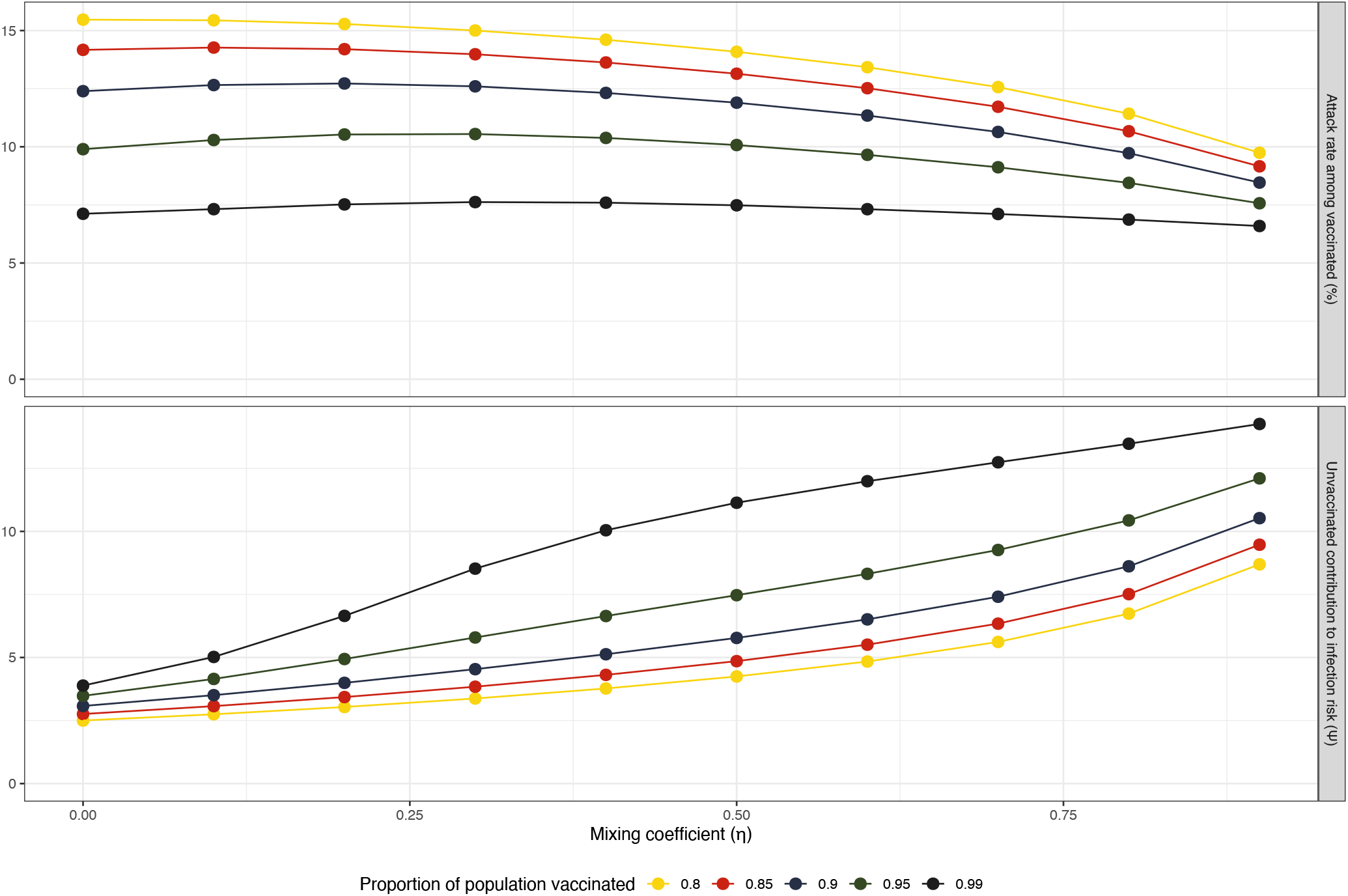
Impact of mixing between vaccinated and unvaccinated populations on contribution to risk and final epidemic size with increasing population vaccination coverage. Increasing population vaccination coverage decreases the attack rate among vaccinated individuals and further increases the relative contribution to risk in vaccinated individuals by the unvaccinated at any level of assortativity. For any given level of vaccination coverage, increasing assortativity decreases the attack rate among the vaccinated but increases the relative contribution to risk in vaccinated individuals by the unvaccinated.

## Discussion

We use a simple deterministic model to explore the impact of assortative mixing on disease dynamics and contribution to risk in a partially vaccinated population during a pandemic modeled on the current pandemic of SARS-CoV-2. Notwithstanding the model’s simplicity, it provides a graphical representation of the expectation that even with highly effective vaccines, and in the face of widespread vaccination, a substantial fraction of new cases can be expected to occur in vaccinated individuals, such that rates, rather than absolute numbers, represent the appropriate metric for presenting vaccination impacts. However, we find that the degree to which people differentially interact with others who are like themselves is likely to have an important impact on disease dynamics, and on risk in individuals who choose to get vaccinated.

Vaccinated individuals were, as expected, at markedly lower risk of infection during the epidemic wave, but when random mixing with unvaccinated individuals occurred, they decreased attack rates in the unvaccinated, serving as a buffer to transmission. As populations became more separate with progressively increasing like-with-like mixing, final epidemic sizes in vaccinated individuals declined, but rose in unvaccinated populations due to the loss of buffering via interaction with vaccinated individuals. Many opponents of vaccine mandates have framed vaccine adoption as a matter of individual choice. However, we demonstrate here that the choices made by individuals who forgo vaccination contribute disproportionately to risk among those who do.

Increased mixing between vaccinated and unvaccinated groups increased final epidemic size among vaccinated individuals; conversely, more assortative mixing decreases final epidemic size among the vaccinated but resulted in enhancement of the degree to which risk among vaccinated individuals can be attributed to unvaccinated individuals. The fact that this excess contribution to risk cannot be mitigated by separating groups undermines the assertion that vaccine choice is best left to the individual, and supports strong public actions aimed at enhancing vaccine uptake and limiting access to public spaces for unvaccinated individuals, as risk cannot be considered “self-regarding”(31). There is ample precedent for public health regulation that protects the wider community from acquisition of communicable diseases, even if this protection comes at a cost of individual freedom (32, 33). As noted above, the use of legal and regulatory tools for the prevention of behaviors and practices that create risk for the wider public also extend beyond communicable infectious diseases, such as statutes that limit indoor cigarette smoking (21-23).

In the context of immune evasion seen with the newly emerged Omicron variant, we find that like-with-like mixing is less protective when vaccine effectiveness is low. This finding underlines the dynamic nature of the pandemic, and the degree to which policies should not be set in stone but need to evolve in a thoughtful manner as the nature of the disease, and the protective effects of vaccines, evolve. Boosting with mRNA vaccines appears to restore vaccine effectiveness against Omicron (5), and it is likely that the higher vaccine effectiveness estimates used in our model will again be relevant to public policy as booster campaigns are scaled up in Canada and elsewhere. The robustness of our findings in the face of wide-ranging sensitivity analysis will allow this work to be applied in future, when new variants arise and as new vaccine formulations become available.

The simplicity of our model is both a strength (as it provides a system that is transparent and easily modified to explore the impact of uncertainty) and a weakness, because it does not precisely simulate a real-world pandemic process in all its complexity. For instance, we modeled vaccine effectiveness against infection but not the additional benefits of vaccination for preventing severe illness. Despite reduced protection against infection by the Omicron variant, vaccinated individuals, including those who have not received third vaccine doses, have continued to receive strong protection against hospitalization and death from SARS-CoV-2 infection (34, 35). This means that acceptance of vaccination is a means of ensuring that greater healthcare capacity is available for those with other illnesses. For example, in Ontario, ICU capacity for COVID-19 cases has been created by cancelling elective surgeries for cancer and cardiac disease, resulting in extensive backlogs (36). By contributing to these backlogs, unvaccinated individuals are creating a risk that those around them may not be able to obtain the care they need, and consequently the risk they create cannot be considered self-regarding. While this benefit is not captured by a simple model focussed on transmission, one advantage of models such as ours is that they provide a ready platform for layering on increasing complexity, so our model can be adapted or expanded to consider impacts on the health system, or to incorporate additional structural elements or alternate assumptions. We have also likely underestimated vaccine benefit in this model, as we have not attempted to capture the impact of vaccines on prevention of forward transmission by vaccinated, infected individuals; this effect appears to be substantial (37). While our focus here is on a pandemic that now appears to be waning, it is unlikely that SARS-CoV-2 will be eliminated, and the findings we present here will be relevant to likely future seasonal SARS-CoV-2 epidemics or in the face of emerging variants.

In summary, this mathematical model demonstrates that while risk associated with avoiding vaccination during a virulent pandemic accrues chiefly to the unvaccinated, the choices of these individuals are likely to impact the health and safety of vaccinated individuals in a manner disproportionate to the fraction of unvaccinated individuals in the population. Risk among unvaccinated individuals cannot be considered self-regarding, and considerations around equity and justice for individuals who do choose to be vaccinated, as well as those who choose not to be, need to be considered in the formulation of vaccination policy.

**Table 1.** Model Parameters.

| Parameter description | Symbol | Value | Plausible<br>Range | Reference |
| --- | --- | --- | --- | --- |
| Probability of transmission per contact<br>multiplied by contacts per year | $\beta$ | 437 | 164-728 | Calculated |
| Rate of recovery from infection (per year) | $\gamma$ | 73 | 41-91 | (38) |
| Basic reproduction number ($R_0$) | $R_0$ | 6 | 4-8 | (3, 7, 28) |
| Mixing between subpopulations (0 =<br>random, 1 = assortative) | $\eta$ | 0.5 | 0-0.9 | Assumption,<br>approach based on<br>(13) |
| Proportion vaccinated | $P_v$ | 0.8 | --- | (39) |
| Vaccine effectiveness | VE | 0.8 | 0.4-0.8 | (3, 7, 40) |
| Population (N) | N | 10,000,000 | --- | (41) |
| Baseline immunity in unvaccinated |  | 0.2 | --- | Assumption |

## Technical appendix

We constructed a simple Susceptible-Infectious-Removed model of a respiratory viral disease, as in (26). The model was subdivided into two connected sub-populations: vaccinated individuals and unvaccinated individuals. A schematic “stock and flow” diagram of the model is presented below. The model is a closed system without births or deaths. Individuals are susceptible (S), infectious (I) or immune (R), and are assigned to a vaccinated or unvaccinated subpopulation, denoted by subscript “v” or “u”, respectively. Force of infection is defined by mixing within and between groups under varying assumptions about assortativity. Here β represents the product of contact rate in a group times infection probability following effective contact. The fraction of contacts among individuals in the i^th^ group (e.g., vaccinated or unvaccinated), with those in the j^th^ group is denoted f_ij_. The rate of recovery is denoted γ. The model assumes durable immunity.

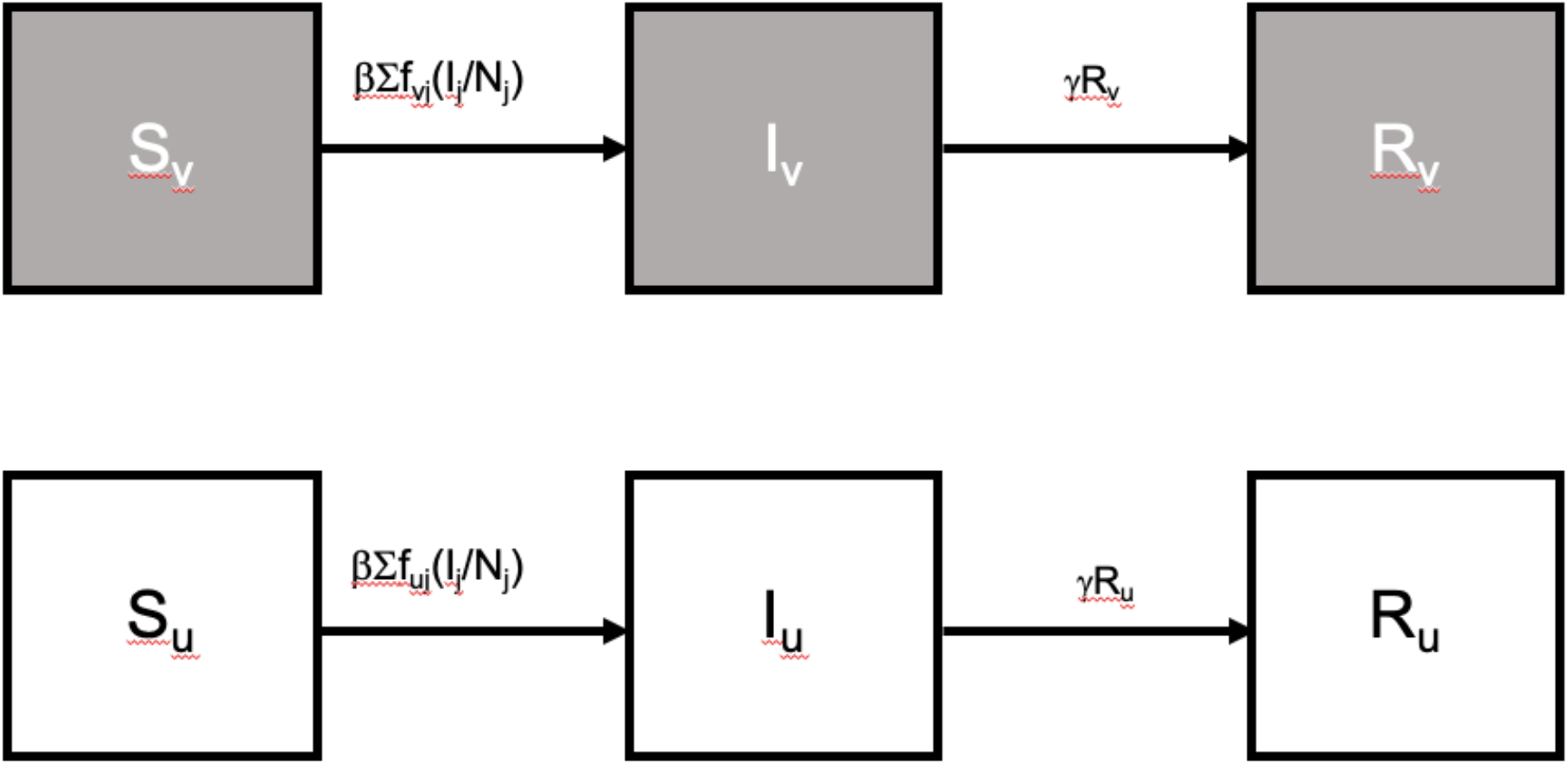

The model is governed by the following ordinary differential equations, where *i* represents the vaccination status of the group, and *j* represents vaccination status of contacts.

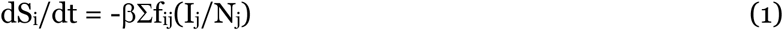

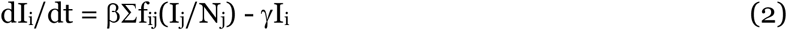

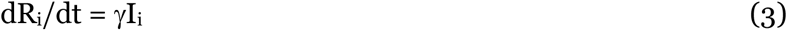

Here β represents the product of contacts per unit time and probability of transmission per contact, which is considered equivalent in vaccinated and unvaccinated individuals, and estimated as R_0_/D, where D is the duration of infectivity and R_0_ is the basic reproduction number. In order to capture non-random mixing between vaccinated and unvaccinated sub-populations, we modeled frequency-dependent transmission, with assortativity modeled using the approach described by Garnett and Anderson (13), which is based on earlier work by Blower and McLean (42), and Gupta et al. (43), and which Blower and McLean refer to as “process models”, to denote the preference inherent in this approach.

Under an assumption of random mixing, the probability of interactions between individuals in different population groups are proportionate to the product of group sizes; if 10% of the population is unvaccinated, under an assumption of random mixing, we would expect 1% of all interactions in the population to involve contacts between unvaccinated individuals (i.e., 0.10 × 0.10 = 0.01). By contrast, we could construct a mixing matrix to describe complete assortativity, in which individuals only interacted with those within their own group; the trace of such a matrix would sum to 1, while the anti-trace would have a sum of zero, as there are no interactions with individuals from other groups. To refer to the example above, under complete assortativity, 10% of all interactions would be interactions of unvaccinated individuals with other unvaccinated individuals, and 90% of interactions would be between vaccinated individuals. The trace of this matrix (which is an identity matrix) would sum to 1 (i.e., 0.9 + 0.1 = 1).

The method described by Garnett and Anderson takes complete assortativity as a starting point, such that the probability of contact between individuals in different groups is represented by the trace of the identity matrix, and the fraction of interactions that occur within the i^th^ group would simply be the fraction of individuals in the i^th^ group (e.g., 10% or 90%, in the example above. Garnett and Anderson denote this probability δ_ij_. In other words, the probability of within group mixing under complete assortativity is:

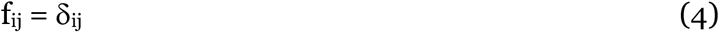

They then redistribute contacts from within-group to between-group contacts based on the propensity towards random mixing in the population, which we denote in our model as η. When we have a matrix of proportions that sum to 1, with p_i_ representing the proportion of individuals in the i^th^ group, and p_j_ (or 1-p_i_ if there are only 2 groups, as here) representing the proportion of contacts in in the jth group, the trace of the matrix is simply Σp_ij_. Our contact patterns are now characterized as:

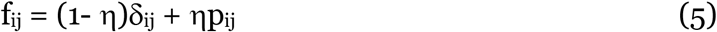

When η is closer to 0 mixing is closer to random, while values closer to 1 represent extreme assortativity. This approach ensures symmetry in contact matrices, such that the number of contacts between vaccinated and unvaccinated individuals is the same, regardless of directionality of contact.

## Data Availability

Data and model are available at Figshare via DOI 10.6084/m9.figshare.15189576

https://figshare.com/articles/dataset/Simple_2-Patch_Vaccine_Model/15189576

## Notes

**Funding statement:** The research was supported by a grant to DNF from the Canadians Institutes for Health Research (2019 COVID-19 rapid researching funding OV4-170360). The funder had no direct role in this work.

### Competing Interest Statement

DNF has served on advisory boards related to influenza and SARS-CoV-2 vaccines for Seqirus, Pfizer, Astrazeneca and Sanofi-Pasteur Vaccines, and has served as a legal expert on issues related to COVID-19 epidemiology for the Elementary Teachers Federation of Ontario and the Registered Nurses Association of Ontario. ART was employed by the Public Health Agency of Canada when the research was conducted. The work does not represent the views of the Public Health Agency of Canada. AA has no competing interests to declare.

### Funding Statement

Canadian Institutes for Health Research

### Summary of Updates

Revised after peer review by Canadian Medical Association Journal.

